# Adaptive Multimodal Fusion in Radiology: Dynamic Balancing of Visual Findings and Clinical Context

**DOI:** 10.64898/2026.09.04.26362226

**Authors:** Carlos Moreno García, Jacinto Mata Vázquez, Victoria Pachón Álvarez

## Abstract

The automatic detection of thoracic pathologies remains a challenge when the visual evidence present in the chest X-ray (CXR) is subtle, ambiguous, or practically imperceptible, and the available textual information is limited, especially in the early phases of patient care. To address this limitation, in this work a clinically motivated subset of MIMIC-CXR is taken as the study set, composed of four diseases associated with dyspnea presentations in the emergency department (heart failure, pulmonary infection, COPD/asthma, and pulmonary embolism), along with a control class. This selection allows evaluating the multimodal fusion of visual and textual information in a clinically diverse set where the contribution of each modality can vary depending on the considered pathology and the quality of the available information.

In this work, a comparative study of different combinations of visual and textual encoders is presented, with the objective of identifying the most suitable configuration for multimodal fusion. Based on this evaluation, we propose *Neural Gated Fusion*, a GMU-based architecture that incorporates an adapted Gated Multimodal Unit to regulate the contribution of visual and textual representations, in contrast to strategies based on static feature concatenation or late decision-level fusion. This architecture uses a neural gate to dynamically regulate the contribution of the visual representation of the radiograph and the textual representation of the clinical report before the multi-label classification stage. The experiments conducted on 25,245 multimodal pairs show that the ViT + ClinicalBERT combination with modulated fusion achieves a macro ROC-AUC of 0.8545 and a macro F1-score of 0.6528, outperforming unimodal models, decision-level fusion, and static early fusion. The per-class analysis suggests that the designed fusion strategy especially improves performance in pathologies with limited visual evidence, such as pulmonary embolism, without compromising the detection of normal cases. These results suggest that the dynamic regulation between image and clinical context is an effective strategy for improving the robustness of multimodal thoracic pathology detection systems.

**Author summary:** Computer-aided diagnosis systems typically rely only on visual data, like chest X-rays, to detect diseases. However, for conditions with subtle or overlapping visual signs (e.g., pulmonary embolism or heart failure), radiologists also rely heavily on patient clinical notes. Our study bridges this gap by creating an AI framework that mimics this real-world diagnostic process. We evaluated multiple visual and text analysis models to find the optimal combination. We then introduced ”Neural Gated Fusion,” an architecture that acts as a smart filter, dynamically deciding whether to prioritize the X-ray image or the clinical report based on the specific case. Tested on over 25,000 multimodal clinical records, our system significantly outperformed traditional single-data models, particularly in diagnosing hard-to-see conditions, without sacrificing accuracy on normal cases. This dynamic approach offers a more transparent and robust tool for clinical decision-making, moving artificial intelligence closer to the holistic reasoning used by medical professionals.

## Introduction

Chest X-rays (CXR) are the most widely requested imaging tests worldwide for the triage and diagnosis of thoracic pathologies [1]. Over the last decade, Deep Learning-based Computer-Aided Diagnosis (CAD) systems have demonstrated a performance comparable to that of human experts [2, 3]. However, most of these systems operate under a unimodal paradigm, exclusively analyzing the visual information from the pixels [4].

Despite their statistical success, purely visual models face a critical limitation: the lack of direct structural evidence in certain lung conditions [5]. Several pathologies, such as heart failure, diffuse Pulmonary Infections, or implicitly diagnosed conditions like pulmonary embolism, can show overlapping or imperceptible radiological patterns in standard non-contrast X-rays [6]. In real clinical practice, radiologists do not interpret an image in isolation; their diagnosis is strongly guided by the patient’s clinical information [7]. The lack of this context in traditional AI models not only limits their accuracy but often leads to spurious or clinically implausible predictions [8].

Given the evidence that simply adding data does not always guarantee better performance, this study is designed to address a specific question regarding how multimodal fusion operates. We pose the following research questions (RQ):

- **RQ1:** How does modality fusion (clinical texts + medical images) contribute to improving the performance of unimodal models, particularly in pathologies with subtle visual patterns?
- **RQ2:** Can adaptive fusion improve performance by dynamically balancing the contributions of visual and textual features?

To answer these questions, we first present an evaluation of multimodal architectures, systematically comparing combinations of advanced visual models (ViT, ResNet50, VGG16) and clinical language models (BioBERT, ClinicalBERT, Biomed-RoBERTa). Based on this analysis, a modulated fusion model termed *Neural Gated Fusion* is proposed. This design aims to demonstrate that the system simultaneously evaluates both sources by adaptively balancing the image and text representations to support robust classification, without prioritizing either modality by default.

## Related Work

### Computer Vision in Medicine

The use of Convolutional Neural Networks (CNNs) has revolutionized medical diagnosis. Architectures such as ResNet [9], Inception [10], Xception [11], DenseNet [12], and EfficientNet [13] have established the state of the art in image classification. Specifically for chest X-rays, massive datasets like ChestX-ray8 [20], CheXpert [19], and MIMIC-CXR [18] have enabled the training of models with strong generalization capabilities. Recently, the literature has shown a paradigm shift toward architectures based on Vision Transformers (ViTs), which outperform CNNs by capturing long-range global dependencies, something crucial for identifying diffuse pathological patterns in high-resolution X-rays [15, 16].

### Natural Language Processing (NLP)

The analysis of medical reports has evolved from rule-based approaches to recurrent neural networks like LSTM [21] and, more recently, Transformers such as BERT [23]. Domain-specific models adapted to electronic health records (EHRs) and biomedical literature, such as BioBERT [24], ClinicalBERT, and biomedical adaptations of Biomed-RoBERTa, have shown superior performance in extracting clinical entities and understanding context. However, since 2023, the emergence of Large Language Models (LLMs) and Generative AI has redefined the state of the art. Models like BioGPT and Med-PaLM have shown clinical reasoning and summarization capabilities that go beyond simple label extraction, enabling a deeper semantic structuring of the patient’s history [25, 26].

### Multimodal Learning and Fusion Strategies

The integration of vision and language has progressed from joint attention mechanisms in TieNet [32] to the alignment between tokens and visual regions to improve report generation [33]. Furthermore, contrastive learning in models like ConVIRT [27] and CLIP [28] has shown that natural language supervision generates robust visual representations. This approach reduces the reliance on manual labels, achieving expert-level accuracy in pathology detection [29].

Recent research has increasingly focused on medicine-specific Vision-Language Pre-training (VLP), aiming to solve the semantic gap between the global image and local textual details. Recent proposals like GLoRIA [30] and MGCA [31] introduce multi-granular alignment mechanisms. Contemporary studies suggest that these aligned representations are essential for developing general-purpose medical AI systems [8, 34].

Standard approaches often rely on feature concatenation (early/feature fusion) or weighted voting ensembles. However, recent literature highlights the challenge of multimodal training, a phenomenon where noise from an uninformative X-ray ends up severely degrading the accuracy provided by the textual branch. Consequently, the design of adaptive fusion architectures (such as *Gated* mechanisms) has become essential to ensure that different data sources complement each other effectively, preventing the limitations of one modality from penalizing the other.

## Materials and methods

The experimental methodology comprises four main phases: (i) strategic curation of the MIMIC-CXR dataset; (ii) establishment of baselines through the evaluation of unimodal (vision and language) models; (iii) implementation of multimodal fusion strategies; and (iv) development of Neural Gated Fusion, designed to dynamically balance the multimodal information.

### Dataset and Preprocessing

For the experimental setup, we used a subset of the MIMIC-CXR dataset, a large-scale repository of chest X-rays paired with textual radiology reports. This dataset is characterized by high clinical variability, making it an ideal setting for multimodal research.

#### Pathology Selection and Sampling Strategy

The dataset was restricted to a clinically selected diagnostic subset composed of four pathological categories and a control class (healthy patients). The selection of these four diseases is based on a purely clinical criterion: they constitute the most frequent causes in patients presenting to the emergency department with dyspnea (shortness of breath) as the primary symptom.

As illustrated in Fig 1, feature extraction in chest X-rays is complex due to the two-dimensional nature of the scan, which causes the superimposition of bone and soft tissue structures, reducing the contrast of subtle lesions. In this context, selecting these pathologies allows the models to be evaluated across different levels of diagnostic difficulty: from clear radiological abnormalities to conditions with diffuse patterns, where the isolated image is ambiguous and requires the context of the clinical report.

- **Heart failure:** Identified by clear structural signs such as cardiomegaly and pleural effusion, although its radiological patterns tend to overlap with other chest conditions.
- **Pulmonary infection:** Representative of pathologies of the alveolar domain, characterized by diffuse borders and high-density opacities.
- **COPD / Asthma:** Obstructive diseases that require the model to identify subtle structural changes.
- **Pulmonary embolism:** A blood clot that blocks a pulmonary artery. Standard X-rays typically show no visible signs at all, making it the most complex case to detect visually.
- **Control (Normal):** Patients with no significant radiological findings, used to measure baseline specificity.

**Figure 1:**
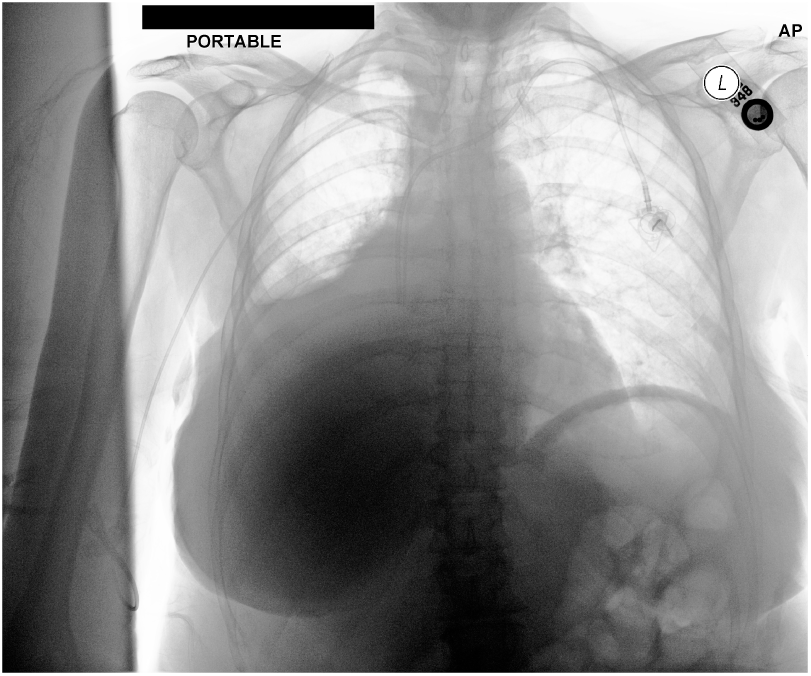
Fig 1 Chest X-ray (CXR) with pulmonary embolism. Frontal image extracted from the MIMIC-CXR dataset, positively labeled for pulmonary embolism.

The final multi-label dataset comprised *N* = 25,245 multimodal pairs. As shown in Fig 2, the resulting class distribution provides a clinically relevant distribution: heart failure (*n* = 11,322), COPD/Asthma (*n* = 7,984), Pulmonary Infection (*n* = 7, 076), pulmonary embolism (*n* = 1,075), and Control (*n* = 7,000). Notably, 7,473 patients presented with two or more concurrent pathologies, which supports the use of a multi-label classification framework and justifies the strong clinical overlap observed in the co-occurrence matrix of the same figure.

**Figure 2:**
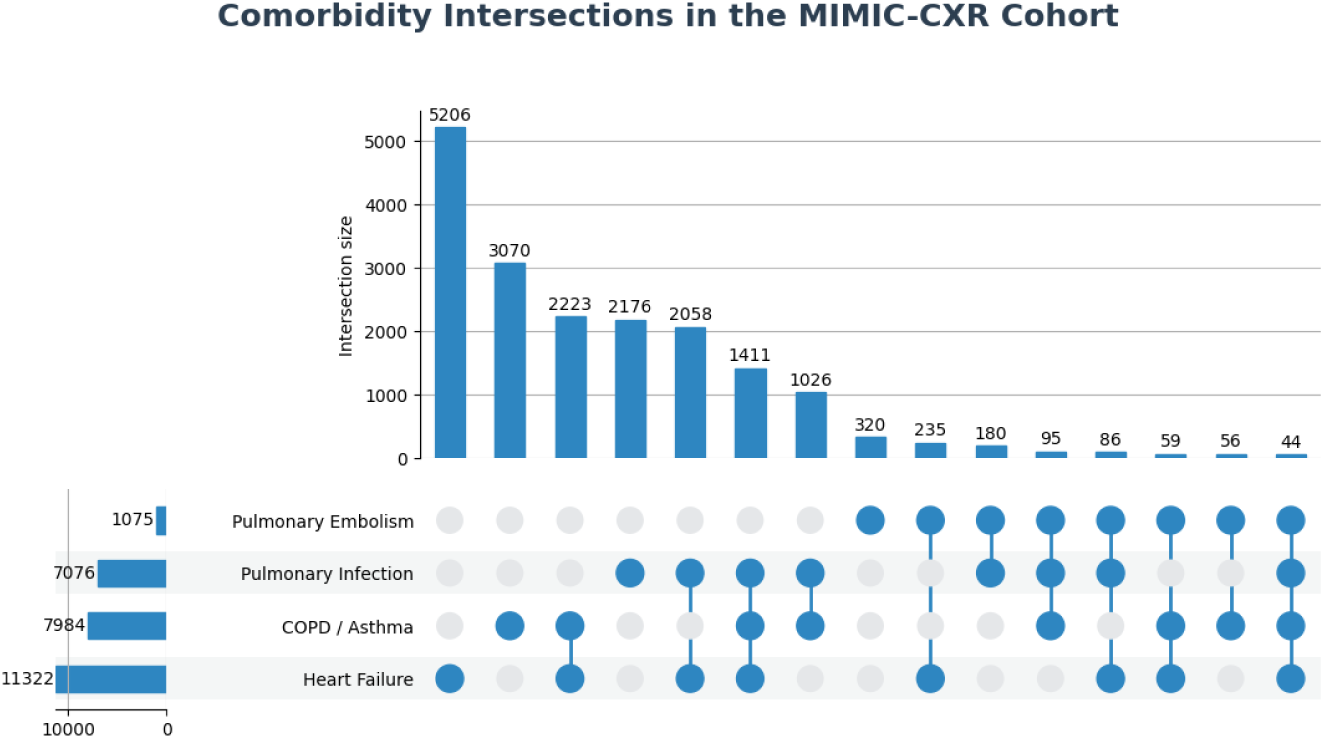
Fig 2 Pathological Overlap in the Dataset. Co-occurrence matrix empirically validating the need for multiple labels (*multi-label*). The diagonal represents the isolated cases for each specific pathology.

#### Data Curation and Filtering Pipeline

To build a robust multimodal dataset, we established a rigorous five-stage extraction and filtering pipeline by cross-referencing data between the MIMIC-IV database, its emergency department module (MIMIC-IV-ED), and MIMIC-CXR, as illustrated in Fig 3.

1. **Overall Hospital Cohort** (*∼***431,000 admissions):** To maximize data capture and represent a realistic clinical distribution, the initial inclusion criteria encompassed all hospital admissions.
2. **Global Diagnostic Labeling:** Patient discharge diagnoses were mapped using ICD-10 codes into the four target respiratory macro-classes. To ensure a strict baseline, the Control group was constructed exclusively from patients presenting no pathological ICD-10 codes in their discharge records, excluding patients who were respiratory-healthy but had other underlying conditions that could introduce radiological noise.
3. **X-ray Alignment (109,636 images):** Admissions were temporally matched with their corresponding imaging studies. The search was strictly limited to frontal views (AP/PA), discarding lateral or technically inadequate scans to ensure the spatial standardization of the visual models.
4. **Text Extraction and Enrichment (MIMIC-IV-ED):** Initially, the textual modality was constructed by extracting the *Indication* sections from the radiology reports. However, since in clinical practice these fields show high sparsity or missing values (e.g., ”N/A”), we implemented a data enrichment strategy. By cross-referencing records with the emergency department module (MIMIC-IV-ED), the reports were augmented by structuring text sequences that integrated demographic data (age, gender) and critical vital signs at the time of triage (heart and respiratory rates, and oxygen saturation, SpO_2_). This decision ensures that the language model receives a continuous and robust physiological context, even when the physician’s narrative is absent. An example of this data-enrichment process is shown in Fig 4.
5. **Strategic Balancing:** To mitigate the severe class imbalance toward the dominant healthy class, selective undersampling was applied exclusively to the Control group, limiting it to 7,000 cases while preserving all pathological instances intact.

**Figure 3:**
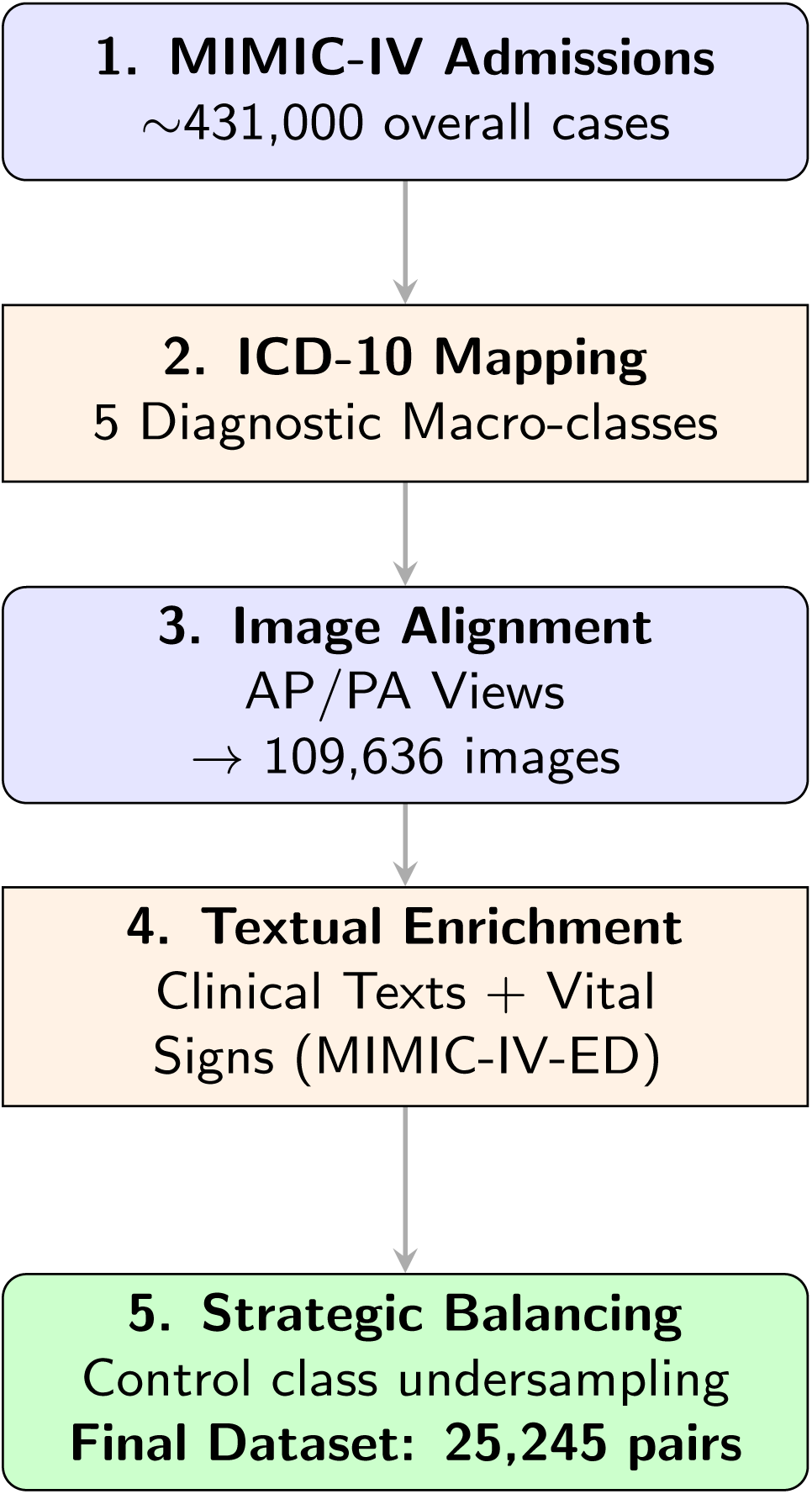
Fig 3 Data Extraction and Curation Pipeline. Sequential flowchart detailing the five-stage filtering process. The methodology not only retrieves the clinical suspicion (*Indications*) but also addresses data sparsity by injecting vital signs from MIMIC-IV-ED.

**Figure 4:**
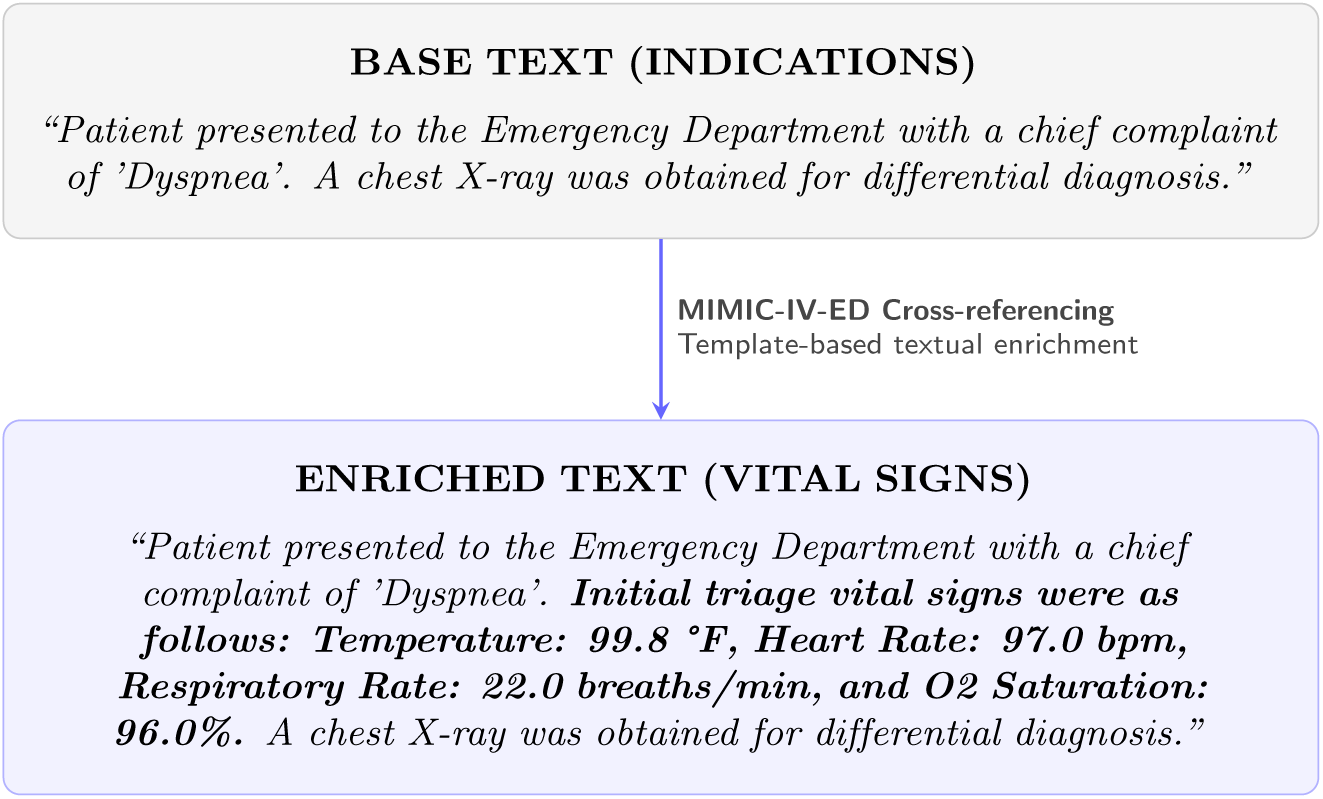
Fig 4 Textual Enrichment Example. Transformation of a baseline clinical report into a dense context input. Triage vital signs extracted from MIMIC-IV-ED are incorporated into the text using a standardised template.

#### Encoding and Transformation

To ensure compatibility between modalities, a rigorous standardization phase was implemented:

- **Pathological Labels:** Modeled as a multi-label classification problem (*y ∈ {*0, 1*}*^5^), allowing for patient comorbidities. Textual labels were processed using a MultiLabelBinarizer.
- **Multimodal Preprocessing:** X-rays were normalized and resized to 224×224 pixels. In parallel, clinical reports were tokenized with a maximum length of 128 tokens. This threshold was established after observing that the maximum text length in our corpus does not exceed 50 words. This way, we ensure the complete capture of the diagnostic context without wasting computational resources processing padding sequences.
- **Label Masking (Concept Blinding):** AI models may memorise explicit keywords to solve a classification problem, bypassing actual medical analysis [38]. To prevent this *data leakage* in our language branch, explicit mentions of the target diseases were masked in the training texts. The models must therefore infer the pathology from the remaining clinical context rather than relying on explicit diagnostic terms [39].

### Unimodal Baselines

Following data preparation, the second phase of our methodology consisted of establishing strong baselines by evaluating the performance of the visual and textual networks separately. The objective of this phase is to measure the individual predictive power of each modality and define the performance boundaries that the multimodal models must surpass.

To establish these baselines, established architectures for each domain will be evaluated independently:

- **Visual Branch (Image-Only):** We evaluate classic convolutional neural networks (ResNet50 and VGG16) to model local spatial features, and global attention-based architectures such as Vision Transformers (ViTs).
- **Textual Branch (Text-Only):** We use pre-trained natural language processing (NLP) models specialized in the medical domain, specifically BioBERT, ClinicalBERT, and Biomed-RoBERTa.

### Multimodal Integration Strategies

To evaluate the interaction between X-rays and clinical reports, we initially implemented the most straightforward approach: aggregating the predictions of the top-performing unimodal models using a Late Fusion (or *Ensemble*) method. However, this approach presents a major structural limitation: by processing the data separately, cross-modal communication during training is prevented, making it impossible to leverage the relationships between image and text [43]. To address this issue, we introduce early fusion, a method where information is unified prior to the classification layer to achieve true cooperation between both sources.

#### Late Fusion

Late fusion constitutes a decision-level approach where the multimodal combination is performed at the final stage of the pipeline. In this framework, the visual and textual models operate entirely independently, processing their respective inputs to yield isolated prediction probabilities (*P_img_* and *P_txt_*).

To obtain the final diagnosis, the system implements a classic *ensemble* via static averaging (50/50 *Soft Voting*). The outputs are combined externally by computing the simple arithmetic mean of the assigned probabilities. However, the fundamental limitation of this architecture is the complete lack of interaction between modalities. Given that the networks operate in isolation from data input to prediction, cross-modal information exchange is impossible. This absence of communication prevents the text and the image from complementing each other, thereby making it more difficult to resolve medical diagnoses.

#### Feature Extraction and Dimensionality Projection

Due to architectural differences among the selected baseline models, the extracted feature vectors present varying sizes. For instance, ResNet50 generates a vector of *d* = 2048, whereas ViT and the clinical language models natively produce *d* = 768. To enable consistent mathematical integration during early fusion, particularly for element-wise operations (*⊙*) the feature dimensions must first be standardised.

As illustrated in Fig 5, this step equalizes the sizes through a projection layer, generating uniform-dimensional vectors (*d* = 768) ready for joint processing.

**Figure 5:**
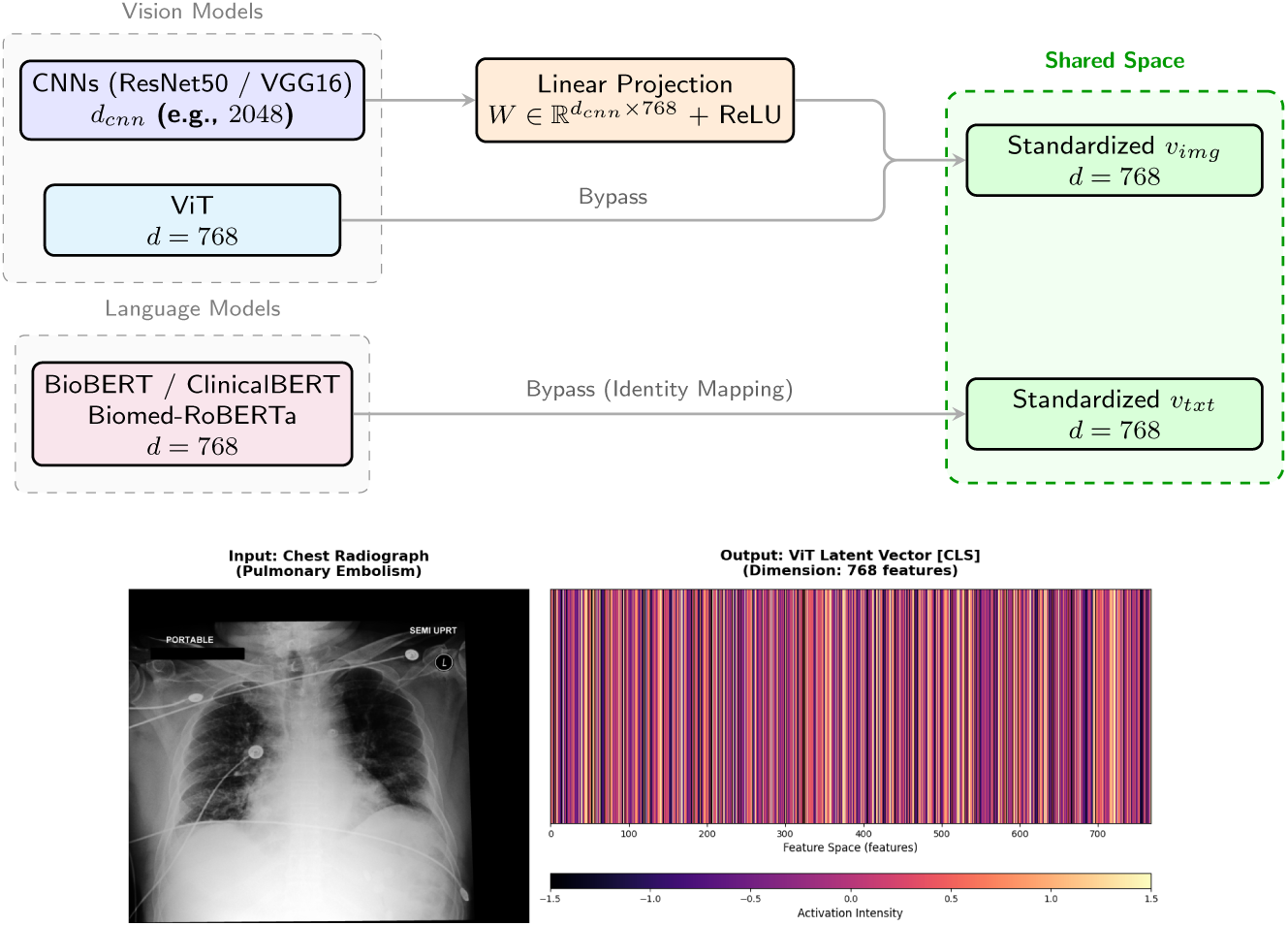
Fig 5 Feature Standardization. **Top:** Dimensionality Mapping Framework. To resolve architectural size discrepancies, high-dimensional representations undergo linear reduction, whereas models natively aligned at *d* = 768 bypass this transformation. **Bottom:** Visual representation of the resulting vector, structured to enable subsequent mathematical operations.

To achieve this standardization, larger outputs (such as those from CNNs) pass through a linear layer that reduces their dimensionality, followed by a ReLU activation function that acts as a filter to eliminate negative values and denoise the signal. Conversely, models already operating at *d* = 768 remain unmodified. This ensures that all features share the same format (*v_img_* and *v_txt_*) as a starting point for the model.

#### Static Early Fusion

Early fusion is a technique that combines features extracted from different modalities at an early stage. Unlike decision-level methods, this approach immediately unifies the representations to feed a single model responsible for performing the final classification. Within this category, feature fusion is implemented. In this static approach, the visual vector (*v_img_*) and the textual vector (*v_txt_*) are concatenated to form a single combined vector, as shown in Fig 6. This new representation directly feeds a dense (*fully connected*) layer to generate the final prediction.

**Figure 6:**
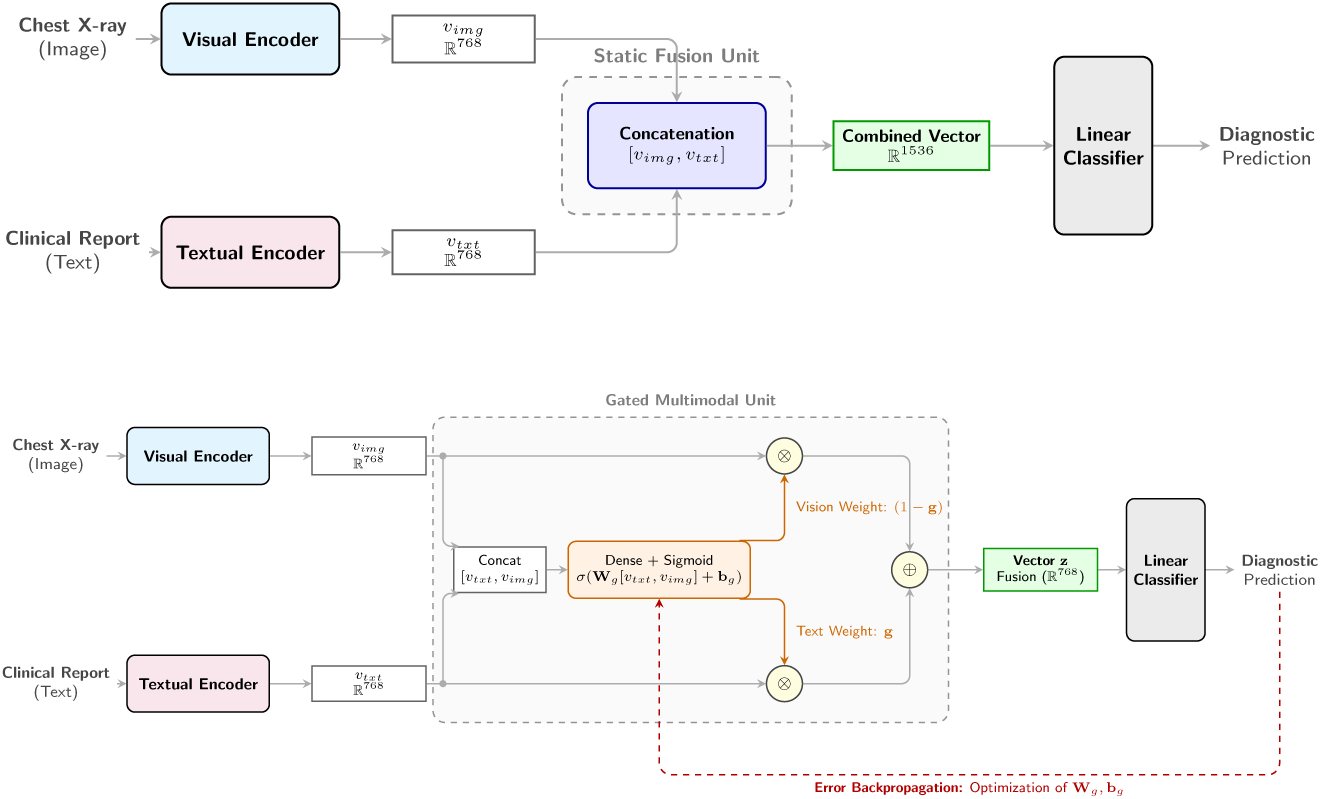
Fig 6 Comparison of early fusion strategies. **Top:** The feature fusion approach groups both vectors statically, doubling the input size to the classifier (R^1536^). **Bottom:** The proposed architecture (*Neural Gated Fusion*) jointly evaluates both modalities in a dense layer to compute a weight vector **g**, which dynamically modulates which visual and textual features should prevail before being summed into a shared space (R^768^).

#### Neural Gated Fusion

The architecture proposed in this work adopts a variant of the Gated Multimodal Unit (*GMU*) introduced by Arevalo et al. [45] as a mechanism for fusing multimodal information. Unlike static feature concatenation, this approach incorporates a neural gate that learns instance-dependent modulation weights. In this work, the GMU is adapted to the radiological domain to combine visual and textual embeddings from pre-trained encoders, allowing the fused representation to be constructed based on the relative contribution of both modalities.

In this architecture, illustrated in Fig 6, the system starts from the embeddings extracted by the unimodal encoders (*v_img_* for the X-ray and *v_txt_* for the report). Unlike approaches that restrict the analysis to a specific multimodal architecture [44], our study proposes a comparative design in which multiple combinations of visual and textual encoders are evaluated. This strategy allows for the empirical selection of the most robust baseline configuration before incorporating the proposed gated fusion mechanism.

Operationally, rather than combining them rigidly, both vectors are concatenated and processed through a linear layer that computes the dynamic weight vector **g**. This vector acts as a gate that determines the relative contribution of the image and text features before combining them into the final representation **z**. The mathematical process is as follows:

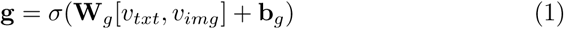

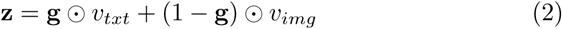

where [*v_txt_, v_img_*] denotes the concatenation of the two embeddings. Given that *v_txt_, v_img_ ∈* R^768^, the concatenation [*v_txt_, v_img_*] yields a 1536-dimensional vector. The GMU layer is parameterized by **W***_g_ ∈* R^768^*^×^*^1536^ and **b***_g_ ∈* R^768^ (internal parameters that are adjusted and optimized as the model trains), and *σ* corresponds to the sigmoid activation function, responsible for bounding the values to the numerical interval [0, 1], producing a vector **g** ∈ R^768^ that assigns a distinct weight to each coordinate of the fused representation.

Through this mathematical formulation, the model establishes an adaptive weighting mechanism. By utilizing element-wise multiplication, the vector **g** evaluates each feature independently to decide how much textual information is retained, while its complement (1 *−* **g**) regulates the contribution of the visual information. The sum of both parts yields the vector **z**. The architecture implements end-to-end learning, while the parameters **W***_g_* and **b***_g_* are learned during training via backpropagation. Once the model is trained, the gate **g** is dynamically computed for each new input both in validation and inference based on the visual and textual characteristics of each case.

Finally, the unified representation **z** feeds the final classifier, a dense (*fully connected*) layer that directly projects the vector onto five output neurons. Its function is to translate this fused information into an independent score for each of the pathologies studied. Since the medical context requires multi-label classification (as a patient may present multiple conditions simultaneously), the model evaluates the probability of each disease separately. Based on these scores, the global error is computed by comparing the model’s predictions with the patient’s actual diagnosis. The backpropagation of this error allows for the simultaneous update of the classifier’s weights and the fusion layer’s parameters (**W***_g_* and **b***_g_*), thereby optimizing the gate based on the success or failure of the diagnosis.

### Implementation Details and Experimental Setup

First, the dataset was split by reserving 20% of the samples for the test set (*test*). The remaining 80% was further subdivided by allocating 90% to training (*train*) and 10% to validation (*valid*). This split ensures that the final evaluation is performed on data unseen by the model.

To ensure the reproducibility of the study, the source code and experimental notebooks are available in a public GitHub repository (https://github.com/CarlosmDTI/Adaptive_Multimodal_Fusion_in_Radiology). The overall implementation was carried out in PyTorch, specifically employing Hugging Face’s transformers library to initialize the models based on the Transformer architecture. All visual and language encoders were initialized from pre-trained weights and fine-tuned for the specific classification task using the data sample.

The training process was configured with a maximum limit of 15 epochs and the AdamW optimizer. The batch size was set to 32 for both the unimodal models and the multimodal approaches. To mitigate overfitting and stabilize learning, a weight decay of 10*^−^*^4^ and a global learning rate of 2 *×* 10*^−^*^5^ were applied. Additionally, an early stopping mechanism was integrated to halt training when no improvements in the macro F1-score were observed on the validation set.

After applying the fusion strategies (Feature and Gated Fusion), the final prediction is performed using a single dense (*fully connected*) layer that directly projects the resulting vector onto 5 output neurons.

Given that a patient may present multiple diseases simultaneously, the problem was formulated as a multi-label classification task, with *y ∈ {*0, 1*}*^5^. The model produces five output logits, one for each pathology considered, and is optimized using the BCEWithLogitsLoss loss function. This function internally combines a sigmoid activation and binary cross-entropy, treating each label as an independent binary decision so as not to impose exclusivity between classes. Unlike a Softmax-based formulation, where classes compete with each other and probabilities are jointly normalized, this approach properly models multi-label scenarios where multiple pathologies can coexist in the same patient.

## Results

In this section, the experimental results are presented in the following sequence. First, the unimodal image and text models were trained and evaluated independently to test the performance of each modality. Next, a late integration strategy at the decision level was evaluated, combining the predictions of the best visual model and the best textual model. Subsequently, early fusion strategies through feature concatenation were analyzed, considering the different combinations between visual and textual encoders. Finally, the proposed modulated fusion architecture was evaluated in order to determine if the incorporation of a neural gate improves the integration of the two information sources. The results are presented following this same experimental sequence and are analyzed using ROC-AUC, F1-score, precision, and recall, employing macro averages to prevent the classes with the highest number of cases from dominating the global evaluation.

### Unimodal Baselines

To establish an initial performance baseline, the two modalities used in the study were evaluated independently: the chest X-ray and the enriched clinical report. This analysis allows quantifies the isolated predictive capacity of each information source and identifies the most suitable encoders for the subsequent multimodal integration strategies.

In the visual branch, the results in Table 1 show that ViT obtains the best overall performance among the evaluated models, with a Macro F1-score of 0.3860 and a Macro ROC-AUC of 0.6957. This behavior suggests that the self-attention mechanism of Vision Transformers is advantageous for capturing broad spatial dependencies in chest X-rays. In contrast, the evaluated convolutional models, ResNet50 and VGG16, present lower performance in aggregate terms, possibly due to their greater reliance on local patterns. Although VGG16 achieves a slightly higher macro recall, this improvement does not translate into a better overall balance between precision and recall; Therefore, ViT was selected as the visual model for the late-fusion experiments.

**Table 1:** Table 1. Performance of the unimodal visual models. Comparison of ResNet50, VGG16, and ViT using only the chest X-ray as input. The F1-score values per diagnostic class and the macro-averaged global metrics used to evaluate the overall behavior of the model are shown.

| Evaluation Metric / Class | Image-Only Baseline Models |  |  |
| --- | --- | --- | --- |
|  | ResNet50 | VGG16 | ViT |
| ***F1-score per Pathology Class*** |  |  |  |
| Heart Failure | 0.4512 | 0.4983 | 0.6480 |
| Pulmonary Infection | 0.2534 | 0.2811 | 0.3448 |
| COPD / Asthma | 0.2981 | 0.3492 | 0.3998 |
| Pulmonary Embolism | 0.0528 | 0.1145 | 0.0275 |
| Control (Normal) | 0.5365 | 0.4964 | 0.5101 |
| ***Aggregated Global Metrics*** |  |  |  |
| **F1-score** | 0.3184 | 0.3479 | **0.3860** |
| **ROC-AUC** | 0.6127 | 0.6438 | **0.6957** |
| **Precision** | 0.3115 | 0.3381 | **0.6509** |
| **Recall** | 0.3342 | **0.3619** | 0.3475 |

It is also observed that even the best visual model presents significant limitations. In particular, ViT obtains a very low F1-score in pulmonary embolism, which highlights the difficulty of detecting pathologies whose radiographic expression may be scarce, indirect, or poorly discriminative. This result indicates that, in the evaluated dataset, isolated visual information does not provide a sufficient discriminative signal for some classes, and therefore, an isolated image may be insufficient for a reliable prediction. This limitation justifies the need to incorporate contextual information from the clinical report.

In the textual branch, the results in Table 2 indicate that ClinicalBERT consistently outperforms BioBERT and Biomed-RoBERTa, achieving a Macro F1-score of 0.5355 and a Macro ROC-AUC of 0.7845. This difference can be explained by the greater alignment between the model’s pre-training and the nature of the texts used in this study, as ClinicalBERT was specifically developed to represent clinical notes. In comparison, more general biomedical models can capture relevant medical terminology, but do not necessarily adapt with the same precision to the style and structure of the clinical texts employed in this work.

**Table 2:** Table 2. Performance of the unimodal textual models. Comparison of BioBERT, Biomed-RoBERTa, and ClinicalBERT using only the textual information from the enriched clinical report. The F1-score values per diagnostic class and the macro-averaged global metrics are presented, with the aim of establishing the predictive capacity of the textual modality.

| Evaluation Metric / Class | Text-Only Baseline Models |  |  |
| --- | --- | --- | --- |
|  | BioBERT | Biomed-RoBERTa | ClinicalBERT |
| *F1-score per Pathology Class* |  |  |  |
| Heart Failure | 0.5210 | 0.5820 | 0.7035 |
| Pulmonary Infection | 0.3805 | 0.4215 | 0.4675 |
| COPD / Asthma | 0.4120 | 0.4530 | 0.4716 |
| Pulmonary Embolism | 0.2505 | 0.3110 | 0.3517 |
| Control (Normal) | 0.5610 | 0.6575 | 0.6832 |
| *Aggregated Global Metrics* |  |  |  |
| **F1-score** | 0.4250 | 0.4850 | **0.5355** |
| **ROC-AUC** | 0.6950 | 0.7320 | **0.7845** |
| **Precision** | 0.4520 | 0.5140 | **0.7373** |
| **Recall** | 0.4010 | 0.4630 | **0.5858** |

Overall, the unimodal baselines support two main conclusions. First, the textual information provides a superior diagnostic performance compared to the isolated image in this experimental scenario. Second, neither modality separately achieves a sufficiently robust performance to predict the pathologies in the dataset. The results of this phase allow the selection of ViT and ClinicalBERT as the reference models for late integration and reinforce the hypothesis that a multimodal strategy can improve the complementarity between chest X-ray images and clinical context.

### Late Fusion

Late fusion was evaluated as a decision-level combination strategy, using the probabilistic outputs of the previously identified best unimodal models: ViT for the visual branch and ClinicalBERT for the textual branch. In this scheme, each model generates its prediction independently, and the combination is performed on the final probabilities through a static average (Soft Voting). This approach allows evaluating whether the aggregation of unimodal decisions is effective in improving overall system performance without modifying the internal architectures specific to each model.

The results in Table 3 show that late fusion clearly improves the performance of both unimodal models. The ensemble achieves a Macro F1-score of 0.5932 and a Macro ROC-AUC of 0.8476, outperforming both ViT and ClinicalBERT separately. This increase indicates that the visual and textual predictions contain complementary information and that their combination at the probability level provides a relevant gain compared to the isolated use of each modality.

**Table 3:** Table 3. Performance Comparison: Isolated Models vs. Late Fusion. Evaluation of the unimodal baselines (ViT and Clinical-BERT) in contrast with the *Late Fusion* strategy. The metrics represent the Macro average to address class imbalance.

| Architecture / Strategy | F1-score | ROC-AUC | Precision | Recall |
| --- | --- | --- | --- | --- |
| *Unimodal Baselines* |  |  |  |  |
| ViT (Image-Only) | 0.3860 | 0.6957 | 0.6509 | 0.3475 |
| ClinicalBERT (Text-Only) | 0.5355 | 0.7845 | 0.7373 | **0.5858** |
| *Late Fusion* (ViT + ClinicalBERT) | **0.5932** | **0.8476** | **0.8223** | 0.5048 |

However, the improvement is not uniform across all metrics. Although late fusion increases the macro F1-score, ROC-AUC, and precision, the macro recall only reaches a value of 0.5048, below the value obtained by ClinicalBERT independently. This result suggests that the static averaging of probabilities may favor a more precise overall prediction, but does not necessarily improve the capacity to retrieve positive cases across all classes. Consequently, decision-level integration constitutes a useful and competitive multimodal baseline, but it is limited for studying more sophisticated cooperation mechanisms between modalities. This justifies the evaluation of early fusion strategies, in which the combination is performed on the feature representations prior to the final classification.

### Static Early Fusion

Static early fusion was evaluated through the direct concatenation of the representations generated by all the visual and textual encoders. Unlike late integration, where each modality first produces an independent prediction, this approach combines both representations before the final classification. In this way, the classifier receives a single multimodal vector and learns the prediction from the joint image and text information.

In this experimental phase, the nine possible combinations between the three considered visual encoders (ResNet50, VGG16, and ViT) and the three evaluated textual encoders (BioBERT, ClinicalBERT, and Biomed-RoBERTa) were evaluated. As shown in Table 4, the ViT + ClinicalBERT combination obtained the best overall performance, with a macro ROC-AUC of 0.8457 and a macro F1-score of 0.6440. This result confirms the trend observed in the unimodal baselines, where ViT obtained the best performance in the visual branch, and ClinicalBERT was the best-performing model in the textual modality.

**Table 4:** Table 4. Evaluation of static early fusion through feature concatenation. Comparison of the nine possible combinations between visual and textual encoders. The configurations are ordered from highest to lowest macro F1-score value.

| Combination | ROC-AUC | F1-score | Precision | Recall |
| --- | --- | --- | --- | --- |
| **ViT + ClinicalBERT** | **0.8457** | **0.6440** | **0.7380** | **0.5969** |
| ResNet50 + Biomed-RoBERTa | 0.8368 | 0.6054 | 0.7447 | 0.5361 |
| ResNet50 + BioBERT | 0.8365 | 0.5949 | 0.7382 | 0.5140 |
| ViT + BioBERT | 0.8347 | 0.5859 | 0.7420 | 0.5231 |
| ResNet50 + ClinicalBERT | 0.8162 | 0.5657 | 0.7443 | 0.4750 |
| VGG16 + Biomed-RoBERTa | 0.8241 | 0.5597 | 0.7350 | 0.4666 |
| VGG16 + ClinicalBERT | 0.8273 | 0.5448 | 0.7429 | 0.4458 |
| ViT + Biomed-RoBERTa | 0.8161 | 0.5326 | 0.7512 | 0.4222 |
| VGG16 + BioBERT | 0.8027 | 0.4834 | 0.7035 | 0.4200 |

The rest of the combinations show that the performance of early fusion depends significantly on the quality of the encoders used in each branch. The configurations based on ClinicalBERT do not always outperform the rest of the alternatives when combined with convolutional visual encoders, which suggests that the effectiveness of the integration does not depend solely on the isolated performance of a modality, but also on the compatibility between the fused representations. In this sense, direct concatenation provides a first form of interaction between image and text, but it does not incorporate any explicit mechanism to regulate the relative contribution of each modality.

The best static early fusion configuration achieves a macro F1-score of 0.6440, representing a relative improvement of 8.56% compared to late integration. However, the macro ROC-AUC remains at a value very similar to that obtained with Soft Voting. These results indicate that feature-level integration improves the overall balance of the classification performance, although the simple concatenation of embeddings does not provide an explicit mechanism to regulate the relative contribution of each modality.

### Neural Gated Fusion

The modulated fusion strategy was evaluated by applying an adapted GMU to the same combinations of visual and textual encoders analyzed in static early fusion. Unlike the direct concatenation of embeddings, this approach introduces a neural gate that calculates dynamic weights to regulate the relative contribution of the visual and textual representation before the final classification. In this way, the model does not simply combine the two modalities, but rather incorporates an explicit weighting mechanism learned during training.

The results in Table 5 show that the ViT + ClinicalBERT combination once again obtains the best overall performance, achieving a macro ROC-AUC of 0.8545 and a macro F1-score of 0.6528. This result represents a relative improvement of 1.37% in macro F1-score compared to the best static early fusion configuration, also based on ViT + ClinicalBERT. Although the gain is moderate, the results indicate that the incorporation of the gate improves the combination of both representations compared to the direct concatenation of features. The behavior observed in the rest of the configurations confirms that the performance of the *Neural Gated Fusion* mechanism depends, to a large extent, on the quality and compatibility of the encoders used.

**Table 5:** Table 5. Performance of Neural Gated Fusion. Comparison of the nine combinations between visual and textual encoders using the proposed dynamic regulation mechanism. The configurations are ordered from highest to lowest macro F1-score value.

| Combination | ROC-AUC | F1-score | Precision | Recall |
| --- | --- | --- | --- | --- |
| **ViT + ClinicalBERT** | **0.8545** | **0.6528** | **0.7268** | **0.6003** |
| ResNet50 + BioBERT | 0.8322 | 0.5926 | 0.7363 | 0.5170 |
| ResNet50 + ClinicalBERT | 0.8291 | 0.5778 | 0.7528 | 0.4910 |
| ResNet50 + Biomed-RoBERTa | 0.8265 | 0.5760 | 0.7463 | 0.4897 |
| ViT + Biomed-RoBERTa | 0.8260 | 0.5663 | 0.7341 | 0.5014 |
| VGG16 + ClinicalBERT | 0.8337 | 0.5552 | 0.7425 | 0.4579 |
| VGG16 + BioBERT | 0.8267 | 0.5504 | 0.7335 | 0.4488 |
| VGG16 + Biomed-RoBERTa | 0.8208 | 0.5460 | 0.7410 | 0.4491 |
| ViT + BioBERT | 0.8143 | 0.5405 | 0.7225 | 0.4361 |

Compared to the previous strategies, modulated fusion provides the best aggregate performance of the entire experimentation. Unlike late integration, it avoids combining decisions already finalized by independent models, and compared to static early fusion, it introduces a learned weighting that allows adjusting the contribution of each modality before the final prediction.

The improvement associated with modulated fusion supports the suitability of incorporating a dynamic regulation mechanism between modalities, capable of adjusting the relative contribution of the visual and textual information.

## Discussion

The global performance comparison, as shown in Table 6, allows for directly answering the research questions of the study.

**Table 6:** Table 6. Global Performance Comparison. Macro-averaged ROC-AUC, F1-score, precision, and recall for the unimodal baselines and the evaluated multimodal integration strategies.

| Model Strategy | ROC-AUC | F1-score | Precision | Recall |
| --- | --- | --- | --- | --- |
| Best Visual (ViT) | 0.6957 | 0.3860 | 0.6509 | 0.3475 |
| Best Textual (ClinicalBERT) | 0.7845 | 0.5355 | 0.7373 | 0.5858 |
| Late Fusion (*Soft Voting*) | 0.8476 | 0.5932 | **0.8223** | 0.5048 |
| Static Early Fusion | 0.8457 | 0.6440 | 0.7380 | 0.5969 |
| **Neural Gated Fusion (Ours)** | **0.8545** | **0.6528** | 0.7268 | **0.6003** |

Regarding the formulation of RQ1, the results confirm that multimodal fusion globally improves performance, given that image and text provide complementary information. Overall, the fusion strategies outperformed the unimodal approaches in terms of global performance, allowing the model to identify pathologies whose visual signs are subtle when analyzed exclusively through the X-ray.

As for RQ2, the global metrics demonstrate that the integration is optimized more effectively through a dynamic regulation mechanism. The proposed *Neural Gated Fusion* architecture achieved the best performance among the evaluated configurations, surpassing the static integration approaches (early and late fusion). The GMU unit dynamically regulates the contribution of the visual information and the clinical context through weights that are optimized based on the error produced during training.

To understand the impact of this mechanism, it is fundamental to analyze the detailed behavior per class of our proposed architecture through its different performance metrics, as detailed in Table 7.

**Table 7:** Table 7. Performance per Class of Neural Gated Fusion. ROC-AUC, F1-score, precision, and recall are reported for each diagnostic category, complementing the macro-averaged evaluation.

| Pathology Class | ROC-AUC | F1-score | Precision | Recall |
| --- | --- | --- | --- | --- |
| Heart Failure | 0.8758 | 0.7610 | 0.7986 | 0.7267 |
| Pulmonary Infection | 0.8334 | 0.5915 | 0.6843 | 0.5208 |
| COPD / Asthma | 0.8428 | 0.6430 | 0.6911 | 0.6011 |
| Pulmonary Embolism | 0.8575 | 0.5640 | 0.7519 | 0.4512 |
| Control (Normal) | 0.8631 | 0.7047 | 0.7080 | 0.7014 |

In pathologies with clear radiographic signs, such as heart failure, the multimodal approach achieves an F1-score of 0.7610, clearly outperforming the 0.6480 obtained by the isolated visual model (ViT) and the 0.7035 of the textual branch (ClinicalBERT).

This improvement is especially notable in pathologies with scarce radiographic expression, such as pulmonary embolism. As reflected in the baselines, the visual model alone performs poorly in this class (F1-score of 0.0275). However, the gating mechanism optimizes the integration of both modalities, raising the F1-score to 0.5640. At the same time, the model preserves its capacity to correctly identify negative cases, achieving an F1-score of 0.7047 in the group of healthy patients (Control).

## Conclusion

In this work, the integration of chest X-rays and clinical reports to detect dyspnea-related pathologies using a sample of MIMIC-CXR data has been analyzed. After systematically evaluating isolated image and text models, static fusion strategies (early and late), and a modulated fusion architecture, three main conclusions are drawn.

First, unimodal approaches based exclusively on the image show significant performance limitations. The most evident case is pulmonary embolism, where the baseline visual model (ViT) barely achieves an F1-score of 0.0275. On the other hand, text models present better predictive behavior, achieving superior performance in these types of cases.

Second, the results show that combining the two branches improves performance compared to the unimodal models. Specifically, the late fusion strategy manages to surpass the metrics obtained in the baseline models. Building on this result, static early fusion further improves performance of the late fusion model by unifying the information at a stage prior to the final decision.

Third, *Neural Gated Fusion* is the most effective strategy of all those evaluated. By unifying the information before classifying, the gate dynamically adjusts the relative weight of the image and the text based on the characteristics of each case. This approach achieves the best performance in the study (macro F1-score of 0.6528), maintaining high precision in pathologies with clear visual signs, such as heart failure, and notably improving detection in the most complex conditions.

## Data Availability

The data underlying the results presented in the study are available from PhysioNet (MIT Laboratory for Computational Physiology). The datasets used in this research (MIMIC-CXR and MIMIC-IV) are restricted-access clinical databases. To protect patient privacy, access requires completing a credentialing process and signing a Data Use Agreement. Qualified researchers may apply for data access directly through PhysioNet at https://physionet.org/content/mimic-cxr/ and https://physionet.org/content/mimiciv/.

https://github.com/CarlosmDTI/Adaptive_Multimodal_Fusion_in_Radiology

https://physionet.org/content/mimiciv/

https://physionet.org/content/mimic-cxr/

